# The vaccine-elicited immunoglobulin profile in milk after COVID-19 mRNA-based vaccination is IgG-dominant and lacks secretory antibodies

**DOI:** 10.1101/2021.03.22.21253831

**Authors:** Alisa Fox, Caroline Norris, Fatima Amanat, Susan Zolla-Pazner, Rebecca L. Powell

## Abstract

The Pfizer/BioNTech and Moderna mRNA-based COVID-19 vaccines are licensed under emergency use authorization, with millions of doses already administered globally [1]. No COVID-19 vaccines are yet under investigation for use in infants or young children. As such, the passive immunity of the antibodies (Abs) provided through milk from a vaccinated person may be one of the only ways to protect this population until pediatric COVID-19 vaccines are licensed. Our early work (as well as an expanded study being published concurrently with this report) examining the milk Ab response after SARS-CoV-2 infection demonstrated that Spike-specific IgA in milk after infection is dominant and highly correlated with a secretory Ab response [2]. Determining if secretory Abs are elicited in milk is critical, as this Ab class is highly stable and resistant to enzymatic degradation in all mucosae - not only in the infant oral/nasal cavity and gut, but in the airways and GI tract as well [3, 4]. Presently, we describe our analysis of the milk Ab response 14 days after completion of an mRNA-based COVID-19 vaccine regimen among 10 individuals. It was evident that unlike the post-infection milk Ab profile, IgG dominates after COVID-19 vaccination. One hundred percent of post-vaccine milk contained significant levels of Spike-specific IgG, with 8/10 samples exhibiting high IgG endpoint titers. Conversely, 6/10 (60%) of post-vaccine samples were positive for Spike specific IgA, with only 1 (10%) exhibiting high IgA endpoint titer. Furthermore, 5/10 (50%) post-vaccine milk samples contained Spike-specific secretory Ab, none of which were found to be high-titer. As our analyses of the immune response in milk to COVID-19 vaccination continues, it will provide a critical opportunity to address huge knowledge gaps, inform the field as to which COVID-19 vaccine, if any, is likely to provide the best milk Ab response, and highlight the need to design improved vaccines with protection of the breastfeeding infant in mind.

## Background

Though pediatric COVID-19 is mild in most cases, ∼10% of infants experience illness requiring advanced care, and even asymptomatic infection can lead to ‘Multisystem Inflammatory Syndrome in Children’ (MIS-C), a rare but potentially deadly inflammatory condition [5-8]. Furthermore, infants and young children can also transmit SARS-CoV-2 to others [9-12]. Clearly, protecting this population from infection remains essential. Several COVID-19 vaccine candidates employing a variety of novel platforms have entered clinical trials over the past 10 months, including the Pfizer/BioNTech and Moderna mRNA-based vaccines, which are now licensed under emergency use authorization, with millions of doses already administered globally [1]. Importantly, none of these COVID-19 vaccines are authorized or currently under investigation for use in infants or young children. As such, the passive immunity of the antibodies (Abs) provided through breastfeeding by a COVID-19-vaccinated mother or milk donor may be one of the only ways to protect this population from SARS-CoV-2 infection and pathology until effective pediatric COVID-19 vaccines are licensed and/or herd immunity is achieved. Mature human milk contains ∼0.6mg/mL total immunoglobulin (Ig), though there is great variation among women sampled [13]. Milk IgG originates predominantly from serum with some local production in specific cases, though IgG comprises only ∼2% of total milk Ab [4]. Approximately 90% of total milk Ab is IgA and ∼8% IgM, nearly all in secretory (s) form (sIgA/sIgM; polymeric Abs complexed to j-chain and secretory component (SC) proteins) [4, 14, 15]. Nearly all sIgA/sIgM derives from the gut-associated lymphoid tissue (GALT), known as the *entero-mammary* link, though there is also homing of B cells from other mucosal-associated lymphoid tissue (MALT), i.e. the respiratory system to the mammary gland. The SC protein is a cleaved segment of the polymeric immunoglobulin receptor (pIgR) which transports this GALT/MALT-derived Abs into the milk.

Early in the pandemic, our team initiated a comprehensive study to assess the SARS-CoV-2-specific Ab response in human milk after COVID-19 recovery. Our early work as well as an expanded study being published concurrently with this report (https://www.medrxiv.org/content/10.1101/2021.03.16.21253731v1) demonstrated that milk from 88% of COVID-19-recovered donors 4-6 weeks after infection was positive for Spike-specific IgA, this IgA response was highly potent, and in 95% of cases, correlated very strongly with a Spike-specific secretory (s) Ab response. This sIgA response is dominant compared to the IgG response, which was present in 75% of samples but of significantly lower titer than sIgA ([2] and newly-submitted data). Determining if secretory Abs are indeed elicited in milk after infection or vaccination is critical, as this Ab class is highly stable and resistant to enzymatic degradation in all mucosae - not only in the infant oral/nasal cavity, but in the airways and GI tract as well [3, 4]. Notably, after 2hrs in the infant stomach, total IgA concentration has been found to decrease by <50%, while IgG concentration decreased by >75%; importantly, though total SC concentration decreased by ∼60%, there was no decrease in the stomach of infants born pre-term (within the first 3 months of life) – a population highly vulnerable to infection [15].

Similarly, we aim to methodically assess the human milk immune response to each of the novel COVID-19 vaccines as such samples become available, as not only are these data critical to public health in the pandemic context, but moreover, there are huge knowledge gaps regarding the human milk immune response to vaccination. Relatively few comprehensive studies exist examining the Ab response in milk after vaccination. The few studies that have examined the milk Ab response after influenza, pertussis, meningococcal and pneumococcal vaccination have generally found specific IgG and/or IgA that tends to mirror the serum Ab response, though none of these studies measured secretory Ab or determined if sIgA was elicited, and data regarding the protective capacity of these milk Abs is conflicting or confounded by the effects of placentally-transferred Ab [16-23]. It is evident from studies in non-human primates (NHPs) that an intramuscular (IM) vaccine may not elicit a robust sIgA response. In a series of experiments with lactating rhesus macaques, an IM DNA prime + IM poxvirus and adenovirus vector boost vaccine regimen was found to elicit specific IgG but virtually no specific IgA in milk [24].

However, NHPs primed IM with poxvirus vector and boosted IM with adjuvanted protein exhibited measurable specific milk IgG and IgA. When alternatively boosted intranasally (IN), specific milk IgA titers were significantly increased [25]. These IN-boosted NHPs were later boosted IM/IN, which produced a markedly high specific milk IgA response [26]. However, it was subsequently determined that this specific IgA was not sIgA, suggesting it did not traffic to milk via the expected route and would be highly susceptible to degradation in the infant mouth and gut. In fact, follow-up study found the IM+IN+IM/IN regimen insufficient to protect breastfeeding infant macaques from oral SHIV acquisition [27]. Notably, there is ∼3-5x less IgA in macaque milk compared to human milk, and macaque milk IgA appears to naturally be comprised of significantly less sIgA compared to human milk; therefore, it is difficult to extrapolate these data onto human vaccination regimens.

The mechanism of for mucosal secretory Ab production after IM vaccination is not fully elucidated, particularly in terms of the potential for sIgA secretion via pIgR rather than passive transfer from serum generating mucosal Ab. It has been suggested that antigen may diffuse from the IM immunization site to local lymph nodes, wherein it is taken up by antigen-presenting cells that initiate a local response followed by migration of these cells and activated lymphocytes to various MALT locales, including Peyer’s patches (PP) in the GALT, which would be critical to the ultimate activation of the entero-mammary pathway and eventual secretion of sIgA in milk [28, 29].

In this first report of our COVID-19 vaccine studies, we describe vaccine-elicited Abs in 10 pairs of milk samples obtained from individual donors 1 day before dose 1, and 14 days after dose 2, of either the Pfizer//BioNTech or Moderna mRNA-based COVID-19 vaccines. Samples were assayed for specific IgA, IgG, and secretory Ab against the full trimeric SARS-CoV-2 Spike protein.

## Methods

### Study participants

Individuals were eligible to have their milk samples included in this analysis if they were lactating, had no history of a suspected or confirmed SARS-CoV-2 infection, and were scheduled to be or had recently been vaccinated with either the Pfizer or Moderna mRNA-based COVID-19 vaccine. If milk samples were determined to be positive for SARS-CoV-2 IgA prior to vaccination, participants were excluded from this analysis. This study was approved by the Institutional Review Board (IRB) at Mount Sinai Hospital (IRB 19-01243). Milk was frozen in participants’ home freezer until samples were picked up and stored at −80°C until Ab testing.

### ELISA

Levels of SARS-CoV-2 Abs in human milk were measured as previously described [2]. Briefly, before Ab testing, milk samples were thawed, centrifuged at 800g for 15 min at room temperature, fat was removed, and supernatant transferred to a new tube. Centrifugation was repeated 2x to ensure removal of all cells and fat. Skimmed acellular milk was aliquoted and frozen at −80°C until testing. Milk was tested in separate assays measuring IgA, IgG, and secretory-type Ab reactivity (the secondary Ab used in this assay is specific for free and bound SC). Half-area 96-well plates were coated with the full trimeric spike protein produced recombinantly as described [30]. Plates were incubated at 4°C overnight, washed in 0.1% Tween 20/PBS (PBS-T), and blocked in PBS-T/3% goat serum/0.5% milk powder for 1h at room temperature. Milk was used undiluted or titrated 4-fold in 1% bovine serum albumin (BSA)/PBS and added to the plate. After 2h incubation at room temperature, plates were washed and incubated for 1h at room temperature with horseradish peroxidase-conjugated goat anti-human-IgA, goat anti-human-IgG (Fisher), or goat anti-human-secretory component (MuBio) diluted in 1% BSA/PBS. Plates were developed with 3,3’,5,5’-Tetramethylbenzidine (TMB) reagent followed by 2N hydrochloric acid (HCl) and read at 450nm on a BioTek Powerwave HT plate reader. Assays were performed in duplicate and repeated 2x.

### Analytical Methods

Control milk samples obtained prior to December 2019 were used previously to establish positive cutoff values for each assay [2]. Milk was defined as positive for the SARS-CoV-2 Abs if OD values measured using undiluted milk from COVID-19-recovered donors were two standard deviations (SD) above the mean ODs obtained from control samples. Endpoint dilution titers were determined from log-transformed titration curves using 4-parameter non-linear regression and an OD cutoff value of 1.0. Endpoint dilution positive cutoff values were determined as above. Mann-Whitney U tests were used to assess significant differences. Correlation analyses were performed using Spearman correlations. All statistical tests were performed in GraphPad Prism, were 2-tailed, and significance level was set at p-values < 0.05.

## Results

Ten pairs of milk samples were obtained from vaccine recipients 1 day before dose 1 and 14 days after dose 2. Six participants had received the Pfizer vaccine, and 4 had received the Moderna vaccine. Milk was used in separate ELISAs measuring IgA, secretory Ab, and IgG binding against recombinant trimeric SARS-CoV-2 Spike [30]. Milk was defined as positive for SARS-CoV-2 Ab if the measured OD or endpoint titer was greater than two standard deviations above the mean value obtained from pre-pandemic control samples, as previously determined for these assays [2]. As part of the screening of participants for this analysis, those with positive Spike-specific IgA in their pre-vaccine sample were excluded; therefore, all pre-vaccine samples in the present study were found to be negative for Spike-specific IgA (Fig. 1a, segmented lines). It was found that 6/10 (60%) of undiluted post-vaccine samples were positive for Spike specific IgA (Fig. 1a, solid lines).

**Figure 1.**
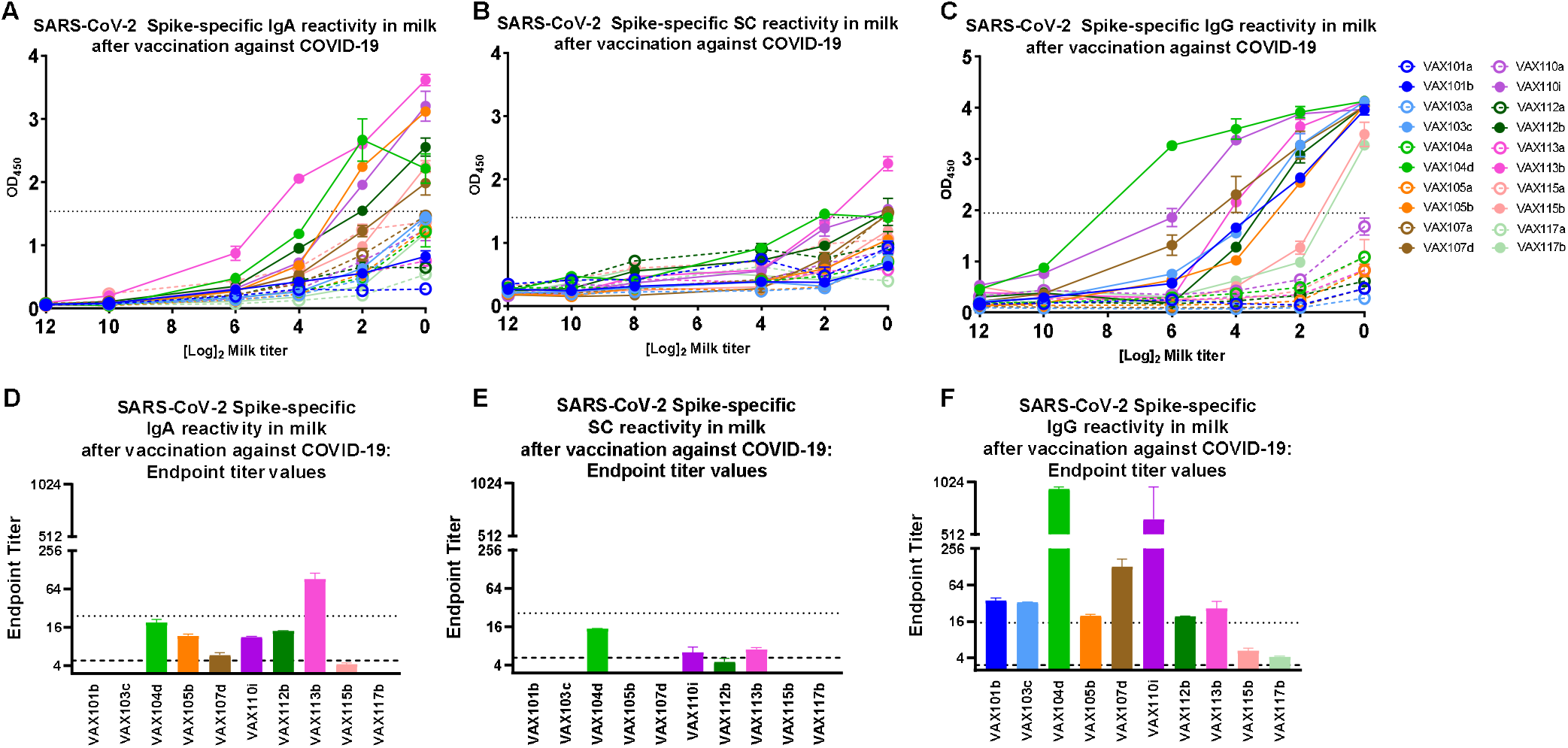
Spike-specific milk Ab profile 14 days post-dose 2 of the Pfizer/BioNTech or Moderna mRNA-based COVID-19 vaccine. Milk was obtained 1 day before dose 1 (open circles/segmented lines) and 14 days post-dose 2 (filled circles/solid lines) from 10 participants. Milk was processed, titrated, and tested by ELISA against the full trimeric Spike for specific IgA (A), secretory Ab (SC: secretory chain) (B), and IgG (C). Dotted lines: positive cutoff values previously determined for each assay as the mean OD of negative control milk samples + 2*SD. (D-F) Endpoint binding titers were calculated for each experiment. Segmented lines: positive cutoff values; dotted lines: 5x positive endpoint cutoff values, designating samples as ‘high-titer’.

As a further measure of antibody affinity/quantity, titrated milk ODs were used to determine endpoint binding titers, finding that of the 6 IgA-positive samples, 5 exhibited positive IgA endpoint binding titers compared to previously-determined cutoff values (83%; Fig. 1d). One sample exhibited Spike-specific IgA endpoint binding ≥5x the positive cutoff, a benchmark we designate as ‘high-titer’ (VAX113; Fig. 1d). Overall, 5/10 (50%) of post-vaccine milk samples contained Spike-specific IgA exhibiting a significant endpoint binding titer.

Next, Spike-specific secretory Ab was measured. It was found that none of the undiluted pre-vaccine samples and 5/10 undiluted post-vaccine samples contained Spike-specific secretory Ab (50%; Fig. 1b). Notably, 4/5 (90%) of these positive samples exhibited binding just at or just above the positive cutoff. Upon titration, 3/5 (60%) of positive samples exhibited significant secretory Ab endpoint binding, with none of these samples exhibiting a high-titer response (Fig. 1e). Overall, 3/10 (30%) of post-vaccine milk samples contained Spike-specific secretory Ab exhibiting a significant endpoint binding titer.

Finally, Spike-specific IgG was measured. It was found that none of the undiluted pre-vaccine samples, and 10/10 (100%) of post-vaccine samples, contained Spike-specific IgG (Fig.1c). Upon titration, 100% of post-vaccine samples exhibited positive endpoint binding titers, with 8/10 samples designated as high-titer (80%; Fig. 1f).

ELISA OD values as well as endpoint binding titers for each assay were compared in separate Spearman correlation analyses (IgG v IgA; IgG v SC; IgA v SC). No correlations were found among any of the parameters measured (data not shown). Additionally, ELISA ODs for each Ab class were compared for milk samples obtained from participants who received the Pfizer vs. Moderna vaccine. No significant differences in values were detected (data not shown).

## Discussion

None of the COVID-19 vaccines currently in clinical trial or authorized for emergency use have been examined for the milk Ab response they elicit. The Pfizer and Moderna COVID-19 vaccines both consist of lipid-encapsulated mRNA delivered IM, both incorporating virtually identical trinucleotide cap analogs, optimized Spike sequences, and N1-methylpseudouridine; however, their lipid carriers differ and the Pfizer vaccine consists of 30ug of RNA while Moderna includes 100ug [31]. In the present study, no differences were detected in milk Ab titers between the groups of participants receiving each vaccine, though numbers in each group were very low and more participants will need to be studied as part of an in-depth longitudinal analysis of each COVID-19 vaccine. IM vaccines have been shown previously to generate mucosal Ab, including Abs in milk, though whether IM vaccination tends to elicit secretory Abs, which would be expected to be the most protective class in a mucosal environment, has generally not been addressed [16-23]. Human milk sIgA is naturally dominant (∼90% of total), and is derived from B cells that transit mainly from the GALT, with some respiratory MALT trafficking as well [4, 32]. Our data investigating the SARS-CoV-2-specific Ab response in milk following infection has demonstrated clearly that this response is robust in most people, and dominated by specific IgA that is largely of the secretory class, while the IgG response is detected in fewer people and is generally of a much lower potency ([2] and concurrently-published data (https://www.medrxiv.org/content/10.1101/2021.03.16.21253731v1)). This natural milk Ab response fits the profile of a classic entero-mammary immunological link, wherein the baby’s oral-nasal cavity is bathed in highly protective secretory Abs generated in response to the pathogens encountered by the mother in the GALT/MALT, a link that evolved to protect infants from deadly mucosal infections [14]. In contrast, our present analysis of COVID-19 mRNA vaccine-induced milk Ab demonstrates a very distinct Ab profile. Among the 10 samples analyzed, the Spike-specific milk Ab profile was consistently IgG-dominant, with all samples exhibiting significant endpoint binding titers, 80% being notably high-titer. Unlike the post-infection response, only 50% and 30% of samples exhibited significant IgA and secretory Ab titers, respectively, which were lower-titer. IgA and secretory Ab data did not correlate, in contrast to our previous findings post-infection, wherein IgA and secretory Ab data was highly positively correlated, demonstrating the IgA was largely secretory class ([2] and concurrently-published data).

Though secretory Abs are vital to the milk Ab defense system, monomeric, non-secretory Abs, originating from serum as well as locally, likely also contribute to the milk Ab defense system, particularly in the case of vaccination [29]. Secretory Ab could ultimately arise in milk following IM vaccination, as will be investigated fully by our follow-up analyses. Importantly, certain vaccine platforms, even if delivered IM, may be more ideal for the elicitation of secretory milk Ab. Depending on the vaccine composition, dose, and adjuvant, antigen may diffuse differentially from the IM immunization site to local lymph nodes, wherein it is taken up by APCs that initiate a local response followed by migration of these cells and activated lymphocytes to various MALT locales, including Peyer’s patches (PP) in the GALT, which would be critical to the ultimate activation of the entero-mammary pathway and eventual secretion of sIgA in milk [28, 29]. Adjuvants as well as immunomodulatory receptors on vectored vaccines may increase and/or modify APC and lymphocyte recruitment, stimulation and trafficking [29]. NHP studies have demonstrated that the vaccine platform and regimen/route is highly significant in terms of the ultimate milk Ab response produced [24-26]. Additionally, the passive transfer of serum Ab into milk should not be ignored, and is likely to also differ among these vaccines based on their differential immunogenicity profiles over time [33-37].

Our continuing studies will ultimately inform the field as to which COVID-19 vaccine, if any, is likely to provide the best protection for breastfeeding infants, so that parents can make an informed decision when such a scenario becomes common. In addition to increasing confidence in these novel vaccines, these data will provide a foundation for a greatly improved understanding of human milk immunology beyond the pandemic, filling large knowledge gaps currently pervading this highly understudied field.

## Data Availability

All data is presented in the manuscript and raw data is available from the corresponding author upon request

## Acknowledgements

As always, we are indebted to the milk donors who make this work possible. This work was supported by the NIH/NIAID.

